# Examining the intersection of child protection and public housing: development, health and justice outcomes using linked administrative data

**DOI:** 10.1101/2021.11.24.21266838

**Authors:** Catia Malvaso, Alicia Montgomerie, Rhiannon Pilkington, Emma Baker, John Lynch

**Affiliations:** BetterStart Child Health and Development Research Group, School of Public Health, Faculty of Health and Medical Sciences, University of Adelaide, Australia; Robinson Research Institute, University of Adelaide, Australia; School of Psychology, Faculty of Health and Medical Sciences, University of Adelaide, Australia; School of Architecture and Built Environment, Faculty of Engineering, Computer and Mathematical Sciences, University of Adelaide, Australia; Population Health Sciences, Bristol Medical School, University of Bristol, United Kingdom, Australia

**Author notes:** **Corresponding author:** Dr C Malvaso, PhD, BetterStart Child Health and Development Research Group, School of Public Health, Faculty of Health and Medical Sciences, University of Adelaide, South Australia, Australia, 5005. **Competing interests statement:** The authors have no competing interests to declare. **Data sharing statement:** The data underlying this article were provided by several Australian State and Commonwealth government agencies under agreements with the researchers led by author J. W. Lynch, SA NT Datalink as the independent linkage authority and multiple ethics committees. Data is only able to be accessed by researchers who have entered into agreements with the Data Custodians and are approved users by the Human Research Ethics Committee. Data can be accessed through an application and approval process administered by the independent data linkage authority, SA NT Datalink. I, the Submitting Author has the right to grant and does grant on behalf of all authors of the Work (as defined in the below author licence), an exclusive licence and/or a non-exclusive licence for contributions from authors who are: i) UK Crown employees; ii) where BMJ has agreed a CC-BY licence shall apply, and/or iii) in accordance with the terms applicable for US Federal Government officers or employees acting as part of their official duties; on a worldwide, perpetual, irrevocable, royalty-free basis to BMJ Publishing Group Ltd (“BMJ”) its licensees and where the relevant Journal is co-owned by BMJ to the co-owners of the Journal, to publish the Work in BMJ Open and any other BMJ products and to exploit all rights, as set out in our <u>licence</u>.

**Keywords:** child maltreatment, child protection, housing, child development

## Abstract

**Objective:** We described development, health and justice system outcomes for children in contact with child protection and public housing.

**Design:** Descriptive analysis of outcomes for children known to child protection who also had contact with public housing drawn from the South Australian (SA) Better Evidence Better Outcomes Linked Data (BEBOLD) platform.

**Setting:** The BEBOLD platform holds linked administrative records collected by government agencies for whole-population successive birth cohorts in South Australia beginning in 1999.

**Participants:** This study included data from birth registrations, perinatal, child protection, public housing, hospital, emergency department, early education, and youth justice for all SA children born 1999-2013 and followed until 2016. The base population notified at least once to child protection was n=67,454.

**Primary outcome measure:** Contact with the public housing system.

**Secondary outcome measures:** hospitalisations and emergency department presentations before age 5, and early education at age 5, and youth justice contact before age 17.

**Results:** More than 60% of children with at least one notification to child protection had contact with public housing, and 60.2% of those known to both systems were known to housing first. Children known to both systems experienced more emergency department and hospitalisation contacts, greater developmental vulnerability and were about 6 times more likely to have youth justice system contact.

**Conclusions:** There is substantial overlap between involvement with child protection and public housing in SA. Those children are more likely to face a life trajectory characterised by greater contact with the health system, greater early life developmental vulnerability, and greater contact with the criminal justice system. Ensuring the highest quality of supportive early life infrastructure for families in public housing may contribute to prevention of contact with child protection and better life trajectories for children.

**Strengths and limitations of this study:**

- This descriptive study provides epidemiological insight into the largely unexplored intersection between child protection and housing systems using whole-of-population linked administrative data
- The findings of this study are based on data drawn from South Australia and may not be directly generalizable to other locations because of different administrative systems
- Despite data being drawn from one Australian jurisdiction, the qualitative relationship between child protection and housing is likely to be similar

## Introduction

Child maltreatment is a worldwide problem and poor outcomes among maltreated children have been well-documented.^1^ Contact with child protection systems is common. In South Australia (SA), 1 in 3 children were reported to child protection by age 18, and 38% were first reported by age five.^2,3^ This cumulative incidence is approximately twice that of asthma.^4^ In Australia, the number of notifications to child protection that were screened-in for review has reached unprecedented levels, with more than 450,000 notifications in 2018-19.^5^ The scale of the child protection problem is consistent with data across Australian jurisdictions,^6,7^ New Zealand,^8^ California,^9^ and the United Kingdom.^10,11^ Extensive reforms within statutory child protection have been recommended by numerous inquiries.^12-15^ A consistent theme is the call for a public health approach to reduce child maltreatment through greater integration of preventive efforts across multiple government agencies. However, the design of siloed ‘incident-based’ information systems (i.e., agencies counting episodes of contact) does not support integration because different agencies may not be able to view their ‘common clients’.^3^

Secure, safe and affordable housing is a basic social determinant of health and is crucial as a stable base for child health and development. Inadequate housing has been suggested as an underlying reason for child protection contact because it reflects resource constraints that limit family capacity to provide adequate care.^16-18^ However, the connection between child protection and housing system contact has been largely unexplored.^19^ We do not know how many children in contact with child protection are also in contact with public housing. Under calls for greater integration across agencies, it is unclear what role the housing system might play in child maltreatment prevention.

This study describes the intersection between the child protection and public housing systems, and subsequent health, developmental and justice system outcomes. First, we documented the number of children in contact with child protection who also had contact with public housing. Second, we identified the proportion of children for whom housing contact preceded child protection notification. Third, we examined perinatal characteristics, emergency department presentations, hospitalisations, early developmental vulnerability, and youth justice system contact for children known to both child protection and housing systems.

## Method

### Data source

The Better Evidence Better Outcomes Linked Data (BEBOLD) platform is a comprehensive linked data platform able to track children’s wellbeing from before birth into early adulthood. It contains deidentified whole-of-population linked administrative data on all South Australian (SA) children born from 1999 onwards. Data were probabilistically linked by an independent linkage agency using demographic characteristics. Australian data linkage systems typically estimate a false linkage rate of 0.1-0.5%.^20,21^

Ethics approval was granted by the SA Department of Health Human Research Ethics Committee (HREC; 377/06/2013; HREC/13/SAH/106), the University of Adelaide HREC (H-185-2011), and the Aboriginal Health Research Ethics Committee (REC2411/9/14). Approval to use these data was also provided by the custodians of each data source.

### Child protection

Information on children who had contact with child protection was obtained from the SA Department for Children Protection (DCP). Children were considered to be in contact with the child protection if they were the subject of at least one report by age 16. In SA, any individual can make a report (known in SA as a notification) to DCP if they suspect on reasonable grounds that a child is, or may be, at risk of child abuse, neglect or harm. Notifications are thus the ‘front-end’ of the child protection system. Notifications are then assessed to determine whether they should be screened-in, and then potentially enter an investigation phase. SA operates under legislation of mandatory reporting for any volunteer or professional who works with children to notify concerns. The base population for this analysis were children notified at least once to child protection was n=67,454.

### Public housing system

Information on children known to the government-funded housing system was obtained from the SA Housing Authority. This Authority collects data on people who have received or applied for government managed housing services and short-term private rental assistance schemes. It does not include schemes funded by the federal government or delivered by non-government agencies. In this study, children were considered to be known to the housing system if they were recorded as living in households who had received at least one of three different types of housing assistance, including: 1) lived in a household receiving short-term private rental assistance; 2) listed as part of a household on a waitlist for public housing; and/or 3) had lived in public housing. These services are offered on a means-tested basis, and because household social and financial circumstances can change it is possible that a child may have experienced all three types of public housing assistance over time.

Short-term private rental assistance provides financial assistance to help secure or maintain a tenancy in the private market, such as bond guarantees, cash bonds, up to two weeks’ rent in advance, rent in arrears (for existing tenancies) and financial assistance for up to three nights’ emergency accommodation. Families who are unable to access suitable housing and who meet income and asset eligibility criteria can apply to live in public housing. They may be placed on a public housing waitlist if they are awaiting approval of an application, awaiting housing becoming available, or awaiting transfer to another property.

### Perinatal characteristics

Perinatal characteristics and demographic information was sourced from the SA Perinatal Statistics Collection. Perinatal data were supplemented and validated by Births Registrations data, which included parental and child demographic information as well as basic clinical birth data. Pregnancy and birth outcome information included: maternal smoking in the second half of pregnancy (yes/no), low birth weight (<2500grams/≥2500grams), preterm gestational age (<37 weeks/≥37 weeks), number of previous births, insufficient antenatal care defined as <7 visits (yes/no), and a postnatal health check at approximately one to four weeks that is universally available (yes/no). Sociodemographic variables included maternal age, maternal marital status (partner/no partner), and parental labour force status (in labour force/not in labour force). Postcode was used to derive a neighbourhood level indicator of sociodemographic disadvantage (Index of Relative Socioeconomic Advantage and Disadvantage; IRSAD)^22^ that included neighbourhood aggregate information on income, education, employment, housing, car ownership, lone parenthood, English proficiency and disability.

### Health, education and justice system outcomes

Contact with adjacent agencies included: emergency department (ED) presentation before age five (yes/no); inpatient public hospital visit (yes/no) before age five; developmental vulnerability at age five on one or more domains (yes/no) and identified as special needs (yes/no) using the Australian Early Development Census (AEDC);^23^ and any contact with the Youth Justice system (yes/no) and admission into custody (yes/no) before age 17.

### Statistical analyses

Data are presented for all SA children born from 1999-2013 for whom child protection and housing data were available. In stage 1 of the analysis, we calculated the proportion of children who had at least one notification to child protection and contact with public housing. The follow-up time from birth to contact with child protection and/or housing was not the same for all children because a child born in 1999 would have had ∼16 years to have contact with these systems, whereas a child born in 2013 would have only had ∼2 years. In stage 2, we calculated the proportion of children who had contact with the child protection and/or the housing system before age five for children born 1999-2010 to ensure that all children had the same follow-up time. In stage 3, we compared sociodemographic and characteristics at birth for children who had contact with the child protection system, ED presentation and inpatient hospital visits before age five, early development at age five, and contact with the Youth Justice system before age 17 (for the 1999 birth cohort only to ensure follow-up to age 17), according to types of housing system contact. For these comparisons, mutually exclusive categories of housing system contact were created to broadly reflect levels of need: 1) no contact with the housing system; 2) children in households which had received short-term private rental assistance but who had never been on a public housing waitlist or had lived in public housing; 3) children in households who had been on a public housing waitlist or who had been in households receiving short-term private rental assistance but had never lived in public housing; and 4) children who had ever lived in public housing before age five. Analyses were conducted in Stata SE version 15.^24^

## Results

Of the children born 1999-2013 who had been notified to child protection at least once (n=67,454), over half (n=40,540; 60.1%) also had contact with public housing. Figure 1 provides a breakdown of public housing contact among children with at least one notification to child protection. The largest proportion (n=33,492; 82.6%) were in a households provided with short-term private rental assistance, followed by those who were ever on a public housing waitlist (n=24,439; 60.3%). A relatively smaller proportion (n=16,078; 39.7%) had ever lived in public housing. However, it was common for children to have experienced combinations of types of housing assistance. As can be seen by the number in the centre of the Venn diagram, just under a quarter of children (n=9,583; 23.6%) had experienced all three types of housing assistance.

**Figure 1.**
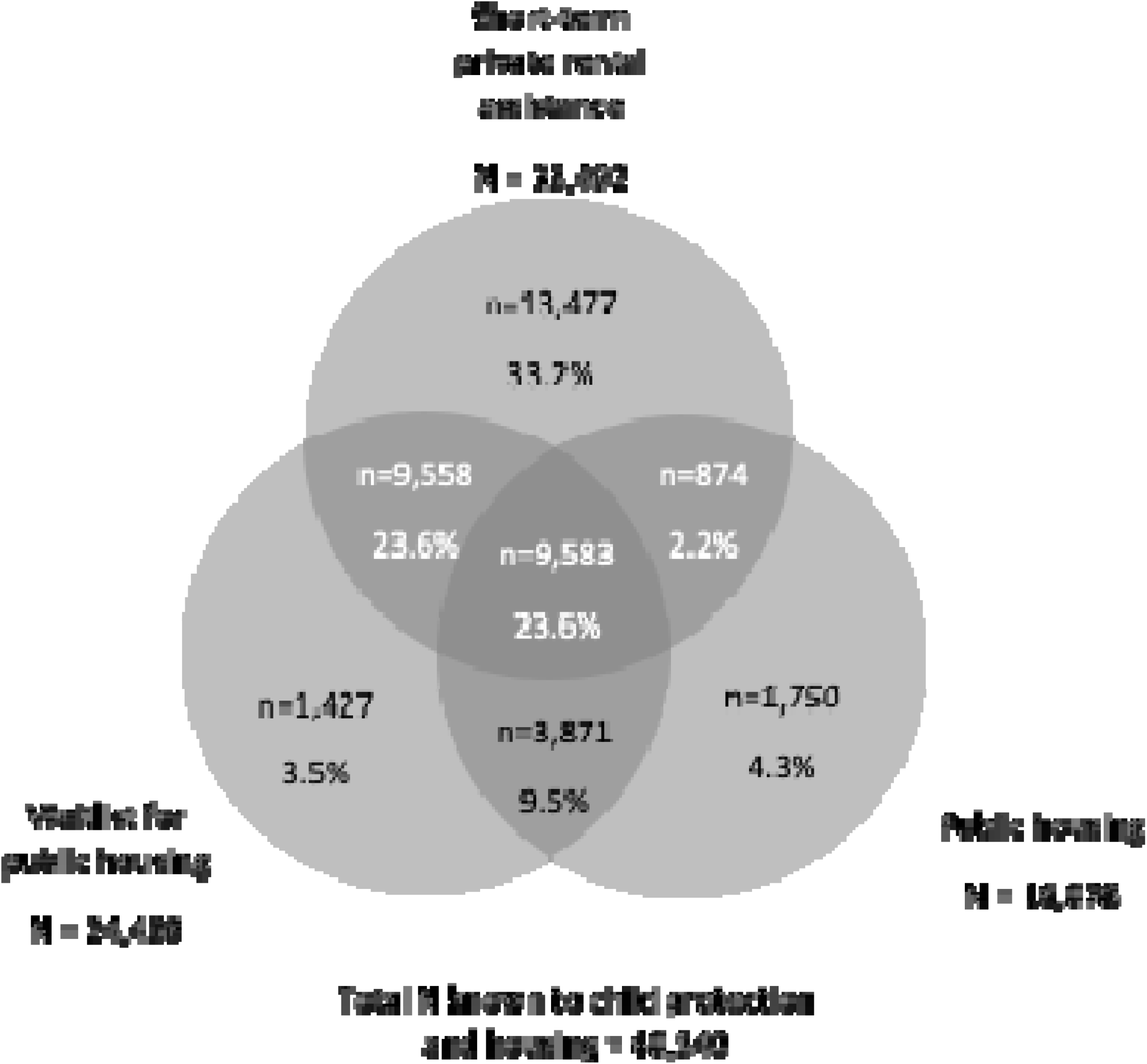
Children born 1999-2013 with at least one child protection notification and different types of housing system contact experienced.

Table 1 shows the number of children with child protection and the housing system contact before age five and includes children born from 1999-2010 to ensure equal follow-up. Before age five, 61.0% of children who were notified at least once to child protection were known to the housing system. Table 1 shows the proportion of children who had contact with housing prior to having contact with child protection before age five. Of those in contact with both systems, the majority (60.2%) were in contact with housing before child protection. This was true for all types of housing system contact. Of the 35,144 children in contact with the child protection system before age five, 12,924 (36.8%) were in contact with housing before child protection contact occurred.

**Table 1:** Timing of child protection and housing system contact before age five for children born 1999-2010.

|  |  |  |  | Different types of housing system contact |  |  |  |  |  |  |  |  |
| --- | --- | --- | --- | --- | --- | --- | --- | --- | --- | --- | --- | --- |
|  | Any housing system contact |  |  | Short-term private rental assistance only<sup>a</sup> |  |  | Listed on a public housing waitlist<sup>b</sup> |  |  | Ever in public housing<sup>c</sup> |  |  |
|  | n | As % of (A) | As % of (B) | n | As % of (A) | As % of (B) | n | As % of (A) | As % of (B) | n | As % of (A) | As % of (B) |
| Notified to child protection (A) | 35,144 |  |  |  |  |  |  |  |  |  |  |  |
| Known to housing and child protection (B) | 21,477 | 61.0 |  | 6,079 | 17.3 |  | 6,241 | 17.8 |  | 9,157 | 26.1 |  |
| Known to housing before child protection | 12,924 | 36.8 | 60.2 | 3,299 | 9.4 | 54.3 | 3,881 | 11.0 | 62.2 | 5,744 | 16.3 | 62.7 |
| Known to housing & child protection at the same time | 73 | 0.2 | 0.3 | 22 | 0.1 | 0.4 | 22 | 0.1 | 0.4 | 29 | 0.1 | 0.3 |
| Known to housing after child protection | 8,490 | 24.1 | 39.5 | 2,758 | 7.8 | 45.4 | 2,338 | 6.7 | 37.5 | 3,384 | 9.6 | 37.0 |

<sup>a</sup> Excludes children who have been in, or on a waitlist for, public housing
<sup>b</sup> Includes children who have been in families receiving short-term private rental assistance but excludes children who have been in public housing
<sup>c</sup> Includes all children who have ever been in public housing

Table 2 shows characteristics measured at birth and outcomes for children born 1999-2010 who had at least one notification to child protection before age five by the type of housing assistance provided. Compared to children with no housing contact, there is a clear pattern of greater social and economic disadvantage for children with the double burden of child protection and public housing contact. Patterns of disadvantage appeared to be greater for children across the different types of housing system support provided, that was meant to broadly reflect levels of housing need.

**Table 2:** Characteristics at birth of children with at least one notification to child protection before age five with different types of housing system contact, born from 1999-2010.

|  | No housing contact<br>(n = 13,667) |  | Short-term private rental assistance only<sup>a</sup><br>(n = 6,079) |  | Listed on a public housing waitlist<sup>b</sup><br>(n = 6,241) |  | Ever in public housing<sup>c</sup><br>(n = 9,157) |  |
| --- | --- | --- | --- | --- | --- | --- | --- | --- |
|  | n | % col | n | % col | n | % col | n | % col |
| **Maternal age** |  |  |  |  |  |  |  |  |
| <19 | 677 | 6.7 | 1,064 | 19.9 | 1,310 | 24.6 | 1,718 | 20.0 |
| 20-24 | 2,053 | 20.4 | 1,880 | 35.2 | 1,724 | 32.4 | 2,577 | 30.0 |
| 25-29 | 2,772 | 27.6 | 1,283 | 24.0 | 1,189 | 22.3 | 2,075 | 24.2 |
| 30-34 | 2,616 | 26.0 | 744 | 13.9 | 704 | 13.2 | 1,378 | 16.1 |
| 35-39 | 1,485 | 14.8 | 305 | 5.7 | 319 | 6.0 | 688 | 8.0 |
| 40+ | 452 | 4.5 | 72 | 1.3 | 81 | 1.5 | 147 | 1.7 |
| **Baby Aboriginal or Torres Strait Islander** | 632 | 6.3 | 370 | 6.9 | 695 | 13.0 | 2,541 | 29.6 |
| **Mother's marital status - no partner** | 2,046 | 20.4 | 1,769 | 33.1 | 2,191 | 41.1 | 3,987 | 46.5 |
| **Mother not in labour force** | 4,899 | 48.9 | 3,397 | 63.7 | 4,027 | 76.0 | 7,276 | 85.3 |
| **Father not in labour force** | 1,556 | 16.2 | 1,173 | 23.5 | 1,816 | 37.5 | 3,941 | 52.1 |
| **Jobless family** | 1,191 | 11.8 | 1,132 | 21.2 | 1,623 | 30.5 | 3,204 | 37.4 |
| **Lived in the most disadvantaged SEIFA quintile** | 3,391 | 33.9 | 2,291 | 42.9 | 2,445 | 45.9 | 4,888 | 57.0 |
| **Mother smoking in pregnancy** | 3,126 | 31.6 | 2,328 | 44.1 | 2,758 | 52.6 | 5,346 | 63.6 |
| **Low birth weight (<2500 grams)** | 896 | 8.9 | 492 | 9.2 | 570 | 10.7 | 1,135 | 13.2 |
| **Preterm birth** | 1,036 | 10.3 | 550 | 10.3 | 625 | 11.8 | 1,187 | 13.9 |
| **Mother number of previous births** |  |  |  |  |  |  |  |  |
| None | 3,685 | 36.7 | 2,255 | 42.3 | 2,116 | 39.8 | 2,437 | 28.5 |
| 1 | 3,108 | 31.0 | 1,555 | 29.1 | 1,476 | 27.7 | 2,128 | 24.9 |
| 2 | 1,761 | 17.6 | 891 | 16.7 | 883 | 16.6 | 1,600 | 18.7 |
| 3 | 863 | 8.6 | 391 | 7.3 | 459 | 8.6 | 1,101 | 11.8 |
| 4+ | 615 | 6.1 | 245 | 4.6 | 385 | 7.3 | 1,387 | 16.2 |
| **Insufficient antenatal care (<7 visits)** | 1,264 | 13.8 | 811 | 16.3 | 1,063 | 21.8 | 2,290 | 29.5 |
| **1 to 4 week health check** | 7,717 | 56.5 | 4,175 | 68.7 | 3,750 | 60.1 | 5,768 | 63.0 |
| **Hospital inpatient before age 5** | 4,961 | 36.3 | 2,801 | 46.1 | 2,765 | 44.3 | 4,635 | 50.6 |
| **Emergency department presentation before age 5** | 4,742 | 34.7 | 2,862 | 47.1 | 2,539 | 40.7 | 3,532 | 38.6 |
| **Developmentally vulnerable at age 5** | 1,035 | 37.4 | 667 | 43.3 | 561 | 46.8 | 1,019 | 56.5 |
| **Identified as special needs at age 5** | 319 | 10.2 | 157 | 9.1 | 154 | 11.2 | 301 | 14.0 |
|  | n | % col | n | % col | n | % col | n | % col |
| **Any Youth Justice involvement up to age 17<sup>d</sup>** | 12 | 1.6 | 8 | 3.6 | 24 | 4.7 | 63 | 10.0 |
| **Entered custody before age 17<sup>d</sup>** | 10 | 1.3 | 5 | 2.2 | 18 | 3.5 | 44 | 7.0 |

<sup>a</sup> Excludes children who had been in, or on a waitlist for, public housing
<sup>b</sup> Includes children who had been in families receiving short-term private rental assistance but excludes children who had been in public housing
<sup>c</sup> Includes all children who had ever been in public housing
<sup>d</sup> Includes children born in 1999 only to ensure follow-up time for Youth Justice to age 17

For example, compared to children notified to child protection but who had no contact with housing, a higher proportion of children who were notified and who also lived in public housing before age five were: born to mothers without a partner (20.4% compared to 46.5%); part of jobless families (11.8% compared to 37.4%); and living in the most disadvantaged areas (33.9% compared to 57.0%). We examined whether children known to both systems had received a postnatal health check that is universally available. Although over half (55.2%) of children known to child protection but who did not have housing system contact did not receive a health check, the proportion was even higher (60-69%) among those who also had contact with the housing system.

Being admitted to hospital and ED presentations was common for all groups. For hospital admissions, this ranged from 44.3% for children known to child protection who had also been listed on a public housing waitlist, to 50.6% for those who ever lived in public housing. For ED presentations, this ranged from 38.6% for children known to child protection who had ever lived in public housing to 47.1% for those who had lived in a household receiving short-term private rental assistance.

The proportion of children identified as developmentally vulnerable on one or more domains at age five differed depending on the type of housing assistance provided. Compared to children notified to child protection but who had no contact with housing, a higher proportion of children who were notified and who ever lived in public housing were developmentally vulnerable (37.4% compared to 56.5%) and a higher proportion were identified as having special needs (10.2% compared to 14.0%).

For the 1999 birth cohort, contact with the Youth Justice system before age 17 also differed across the different types of housing assistance, with 10% of children who were notified and who lived in public housing before age 5 experiencing Youth Justice supervision and 7% entering custody by age 17 (compared to 1.6% and 1.3% who had only been notified). See supplementary material for subsequent birth cohorts.

## Discussion

There is substantial overlap between children known to the child protection and public housing systems in SA. Over half of the children born 1999-2013 who were notified at least once to child protection also had contact with the housing system, and over 60% of those known to both systems were known to housing first. Children known to both systems experienced a greater burden of hospitalisations, emergency department presentations, developmental vulnerability and a higher likelihood of youth justice system contact. There may be potential for the public housing system to be a focus for prevention efforts in child maltreatment given our findings that over a third of all the children who came to the attention of child protection by age five were already receiving some form of housing assistance.

A coordinated service approach from agencies providing housing assistance and those providing family support might better meet the needs of some families rather than services operating in isolation. With ongoing support to ensure basic housing needs are met, families may be better able to engage with other support services. Housing workers could be upskilled to provide the relevant outreach and community connections to support parenting-related needs. Using housing as a conduit to community-based support services may be viewed as less threatening because contact and support are provided while delivering a housing benefit rather than in the context of child protection where parenting practices are scrutinized.

Despite their preventive potential, housing agencies are resource constrained and have failed to keep pace with need with 149,000 households waiting housing allocation in 2019.^25^ A decline in the proportion of public housing stock has occurred alongside housing affordability crisis in Australia putting many households at increased risk of financial stress, which may impact child wellbeing.^26^ Unmet demand for homelessness services is also increasing, with 32.7% of individuals with an identified need being unmet in 2017-18.^27^ Individuals experiencing family violence comprised 40% of specialist homelessness services clients in 2016-17, with more than one-fifth of clients (22%) including children under the age of 10.^28^

Although the findings of this study are based on data drawn from SA and may not be directly generalizable to other locations because of different administrative systems, the qualitative relationship between child protection and housing is likely to be similar.

This paper is the first to demonstrate the large overlap between families known to child protection and public housing systems. It is well known that there are multiple drivers of child maltreatment, including family violence, serious mental health issues, and drug and alcohol abuse and that these often co-occur with poverty and poor housing. Secure, safe and stable housing is a fundamental social determinant of health. This study has shown the added health, developmental and criminal justice burden for the substantial proportion of children experiencing both child protection and housing contact. Ensuring the highest quality of supportive early life infrastructure for families in public housing may contribute to prevention of contact with child protection and better life trajectories for children.

## Data Availability

The data underlying this article were provided by several Australian State and Commonwealth government agencies under agreements with the researchers led by author J. W. Lynch, SA NT Datalink as the independent linkage authority and multiple ethics committees. Data is only able to be accessed by researchers who have entered into agreements with the Data Custodians and are approved users by the Human Research Ethics Committee. Data can be accessed through an application and approval process administered by the independent data linkage authority, SA NT Datalink.

## Author contributions

All authors were involved in study conception and design, interpretation of the data, and drafting of the manuscript. JL, RP, AM and CM were involved in the acquisition of data. AM and CM analysed the data. EB and JL provided critical revision of the article.

